# Personalized, closed-loop deep brain stimulation for chronic pain

**DOI:** 10.1101/2025.08.11.25333010

**Authors:** Prasad Shirvalkar, Ryan Leriche, Jeremy Saal, Jackson Cagle, Jordan Prosky, Isabella Joseph, Ana Shaughnessy, Ashlyn Schmitgen, Joanna Lin, Julian Motzkin, Heather Dawes, Caroline Racine, Andreea Seritan, Kristin K. Sellers, Coralie deHemptinne, Philip A. Starr, Edward F. Chang

**Author notes:** These second authors contributed equally (Ryan Leriche, Jeremy Saal). <u>Corresponding Author:</u> Prasad Shirvalkar MD, PhD University of California, San Francisco Office 671H, Box 3128 1651 4^th^ Street San Francisco, CA 94158.

## Abstract

**Background:** Chronic pain is a major healthcare problem associated with maladaptive brain circuit changes - many patients are unresponsive to all available therapies. Deep brain stimulation (DBS) is a promising treatment, but traditional targets have not proven consistently effective. DBS for pain may be improved by individualizing location and timing of stimulation based on real time brain measurements and patient reports.

**Methods:** To optimize personalized stimulation targets, we first performed a double blind, sham-controlled, intracranial EEG brain mapping trial spanning 10 hospital days, in six participants with refractory neuropathic pain syndromes. Five participants with clinically meaningful pain relief were then implanted with permanent devices capable of brain stimulation and recording. We used ambulatory electrical brain recordings to derive bespoke pain biomarkers using machine learning. Pain biomarkers were used in closedloop DBS algorithms for personalized therapy. After an open-label period, we tested the feasibility and efficacy of closed-loop DBS for pain relief against sham in a double-blind, cross-over trial.

**Results:** Trial intracranial testing revealed multiple brain targets among cortico-striatal-thalamocortical pathways that produced rapid pain relief across participants. We predicted individual pain metrics from both inpatient and ambulatory brain activity with high accuracy; pain biomarkers were incorporated into closed-loop DBS algorithms that also responded to sleep-wake cycles. Personalized, closed-loop DBS was superior to sham, with durability up to 3.5 years.

**Conclusions:** Precision-medicine DBS, with individually optimized brain stimulation targets and closed-loop delivery of stimulation in response to pain biomarkers, is a feasible strategy to treat refractory chronic pain syndromes.

**Trial Registration:** NCT04144972

## INTRODUCTION

Chronic pain affects up to 20% of the US population^1^, but many pain syndromes are refractory to current therapies. Because neuropathic pain syndromes are associated with brain-wide circuit changes^2–5^, deep brain stimulation (DBS) is an appealing strategy to treat refractory cases. Despite over 65 years of human study^6^, pain DBS outcomes have been inconsistent with poor long-term results. Prior DBS attempts have used a one-size-fits-all approach by targeting the same brain regions with continuous stimulation across patients despite interindividual variation in brain anatomy and physiology. Individualizing targets and stimulation parameters of DBS for pain using real-time pain biomarker feedback offers an opportunity to significantly improve clinical outcomes.

Traditional DBS targets for pain relief include the periventricular gray (PVG) and sensory thalamus (VPthal) which play key roles in endogenous opioid release and ascending pain transmission, respectively^7,8^. The majority of prior studies lack blinding or placebo control, with rare exceptions^9,10^. Recent neuroimaging and electrophysiology studies of pain reveal numerous other hubs that may engage descending pain modulation circuits for analgesia^11–14^. These include the anterior cingulate cortex (ACC), insula, ventromedial prefrontal cortex (vmPFC), basal ganglia^13^ and centromedian thalamus (CMThal), which all participate in pain-relevant cortico-striatal-thalamocortical loops^13,15^. Although ACC^16^ and CMThal^17^ DBS have shown variable benefit in case-series, rigorously identifying the best stimulation target for a particular individual has remained elusive^18^.

Invasive brain stimulation with intracranial EEG (iEEG) electrodes can help identify optimal targets for pain relief in each individual. iEEG is commonly used in drug-resistant epilepsy to identify seizure foci and map cortical function^19^. We recently demonstrated the feasibility of using iEEG to identify individualized biomarkers of real-world chronic pain symptom state over many months^20^. By integrating such biomarkers into personalized, adaptive brain stimulation algorithms, we performed an early feasibility trial to assess the safety, efficacy, and durability of individually tailored neurostimulation in response to brain signatures that tracked clinical pain symptoms. Such adaptive brain stimulation has been proposed to potentially avert side effects or the development of habituation to electrical stimulation in patients with major depressive disorder^21^ and Parkinson’s disease^22^.

Here we describe the proof-of-concept treatment of refractory chronic neuropathic pain syndromes using a double-blind and placebo controlled, temporary iEEG stimulation to inform long-term closed-loop DBS. In five participants, we observed acute, rapid pain relief which translated into long-term efficacy after permanent closed-loop DBS for up to 3.5 years. We discovered two novel targets for pain DBS in the basal ganglia, left caudate body and ipsilateral globus pallidus internus. We successfully deployed closed-loop DBS at key cortico-striatal-thalamocortical targets in response to real-time, bespoke neural biomarkers of chronic pain severity and relief. Closed-loop DBS was superior to sham control across participants. We demonstrate the feasibility of closed-loop DBS and long-term efficacy of a precision-medicine approach for brain stimulation for refractory chronic pain.

## RESULTS

### Identifying personalized stimulation targets in a temporary trial

The primary goal of the 10-day, inpatient trial was to identify one optimal target for pain relief in each cerebral hemisphere per participant. Up to 14 candidate sites were tested per hemisphere, based on key nodes identified from the pain DBS and neuroimaging literature (Figure 1).^2,11,12,18,24,26^ We first determined the best stimulation frequency for each site by stimulating at 10, 50 or 100 Hz for 30 seconds assuming that transient stimulation would predict long-term pain relief (Section S5).

**FIGURE 1:**
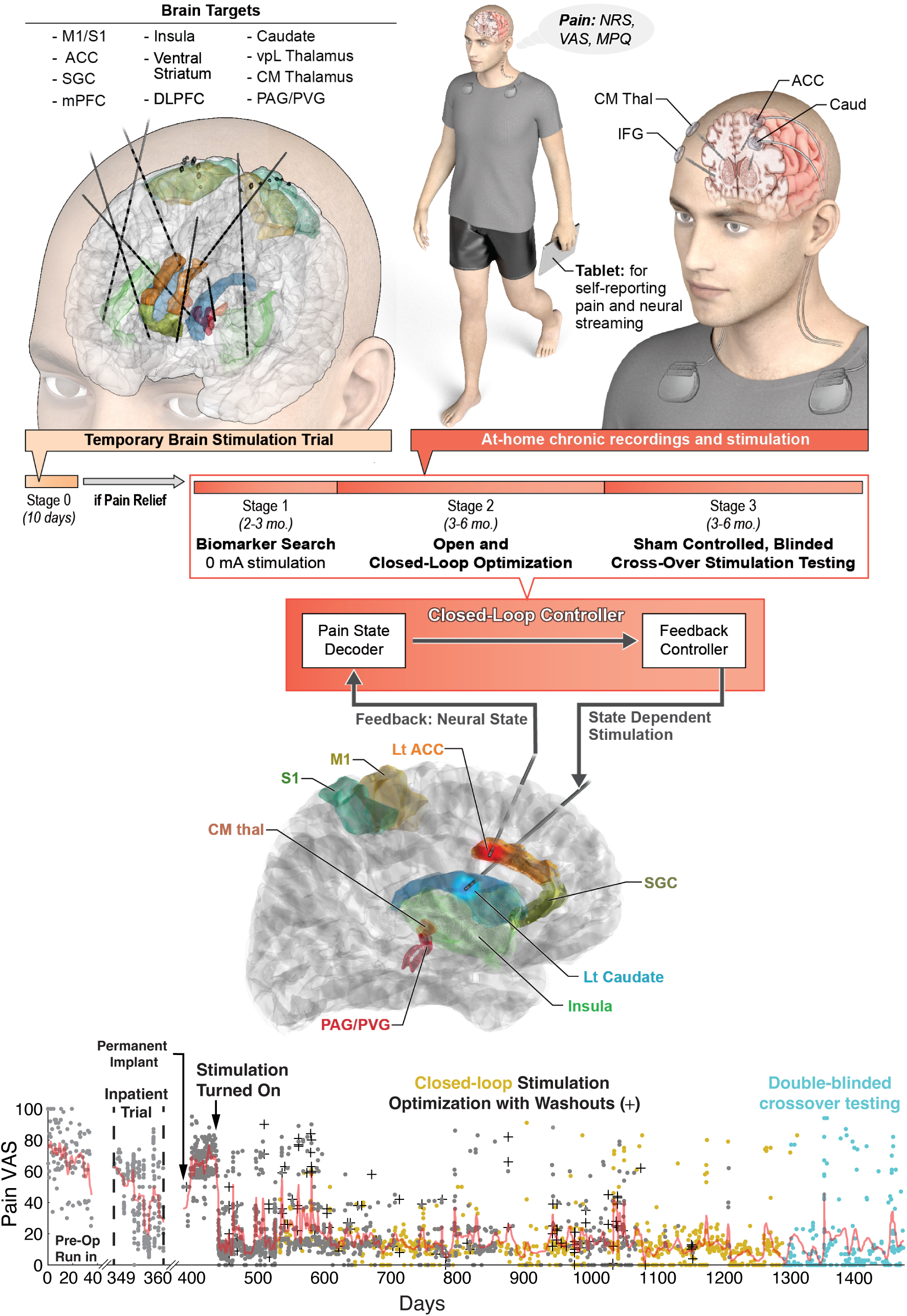
Study overview schematic of precision DBS for chronic pain. Participants underwent a 10 day inpatient stimulation trial where various brain targets in each hemisphere were implanted with sEEG and ECoG electrodes and electrically stimulated with bipolar, biphasic pulses to evaluate for pain relief in comparison to sham (0 mA) stimulation. If participants experienced clinically meaningful pain relief over multiple sessions, they were then permanently implanted with two Summit RC+S DBS systems connected to electrodes targeting the optimal, personalized stimulation and biomarker recording targets (Left Caudate and ACC and Right CMThal and IFG, P3). The RC+S system communicated with a tablet computer to record neural signals and participants self-reported pain symptoms (e.g., NRS, VAS, MPQ) multiple times daily. Pain predictive biomarkers were identified with machine learning decoders. Biomarkers were used to optimize personalized, closed-loop DBS algorithms that could activate stimulation in response to detected high pain states through an embedded feedback controller (brain schematic below). (Bottom panel) Example pain VAS score reports, with moving average (red line) from a participant over 1500 days of trial participation illustrating preoperative run-in, temporary inpatient brain stimulation trial period, followed by permanent DBS implant, open-label (grey) and closed-loop optimization (yellow) with periods of washout (cross), and double-blinded, sham controlled, closed-loop DBS program testing (cyan). (Lt ACC Left anterior cingulate cortex, M1 primary motor cortex, S1 primary somatosensory cortex, CMThal centromedian thalamus, PAG/PVG periaqueductal / periventricular gray, SGC subgenual cingulate, IFG inferior frontal gyrus, Caud caudate, VAS visual analog scale, NRS numerical rating scale, MPQ McGill Pain Questionnaire short form version 2)

Across 5 of 6 participants (P1–P5), multiple targets yielded significant reductions in pain VAS compared with sham, but the magnitude and topography of these responses varied considerably (Figure 2, Tables S3-8). P1 demonstrated significant reductions of up to 30 points VAS from baseline at the most effective targets (U_LVCVS_=24.5 p=0.018, U_RVPN_=168 p=0.016, U_RACC_=101 p=0.011), while P2 achieved larger improvements up to 60 points compared to pre-stimulation baseline scores (U_LCaud_=127 p=7×10^-4^, U_RCMThal_=165.5 p=0.041, U_RACC_=67 p=0.012). P3 also exhibited large significant pain reductions (up to 60, U_LCaud_=955.5 p=0.011, U_RCMThal_=473 p=0.016), whereas P4 showed more modest, nonsignificant reductions of VAS in the 20-point range (U_LACC_=442 p=0.43, U_RCMThal_=287.5 p=0.61). Finally, P5 attained up to 45 points of improvement on average, which frequently corresponded to a 0/100 VAS rating (U_LGPi_=33 p=0.007, U_RCMThal_=234.5 p=0.001). We discovered two novel targets that acutely reduced pain: the globus pallidus internus and the left caudate body (Videos S1-2). Sham stimulation in five participants produced modest analgesia, consistent with a placebo effect. Although targets in P4 did not reach statistical significance, his improved function and ability to hug his wife for the first time in years with LACC and R CMThal DBS were considered meaningful enough to advance to permanent implant. Despite interindividual variability in magnitude and anatomical target, all participants but one experienced clinically meaningful pain reduction with at least one site of DBS per hemisphere.

**FIGURE 2:**
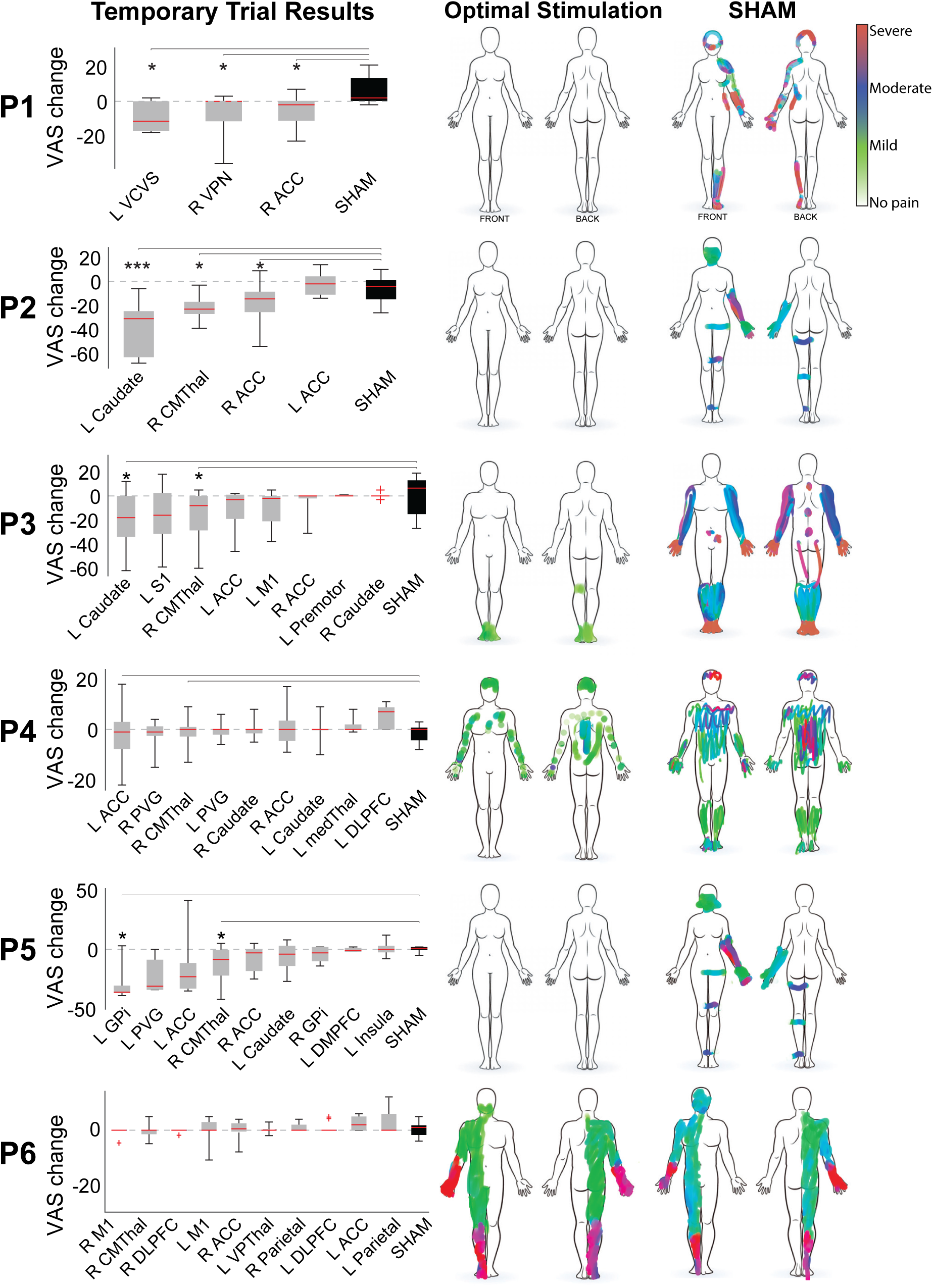
Temporary brain stimulation trial identified different optimal DBS targets across participants 1-6 (P1-6). Each row corresponds to a single participant. Box plots show change in pain VAS from pre stimulation baseline to after double-blinded stimulation at various targets over repeated trials (sorted by median per participant). Red bar indicates median value, grey boxes represent first and third quartiles, and whiskers show full data range. Representative pain body diagrams drawn by participants immediately after optimal stimulation (middle panel) or sham stimulation (right panel) reflect pain location and intensity (color bar). (*p<0.05, ***p<0.001, L left, R right, VCVS = ventral caudate /striatum, VPN/L ventral posterior thalamus, ACC anterior cingulate cortex, CMThal centromedian thalamus, S1 primary somatosensory and M1 primary motor cortex, PVG periventricular gray, DLPFC dorsolateral prefrontal cortex, GPi globus pallidus internus.) P6 did not have any clinical meaningful pain relief so no statistical testing was done.

### Identifying optimal sites for pain biomarkers in temporary trial

In each of five participants, we identified biomarkers of chronic pain by aggregating results across 16 different machine learning models. We trained classification and regression models using iEEG signals from multiple preplanned durations to predict self-reported pain metrics (Figure 3A). Participants provided an average of 266 pain metrics over 10d (range 224-360) including NRS, VAS, unpVAS and reliefVAS (Figure S5.2), each occurring at least 5 minutes away from stimulation to mitigate signal artifacts.

**FIGURE 3:**
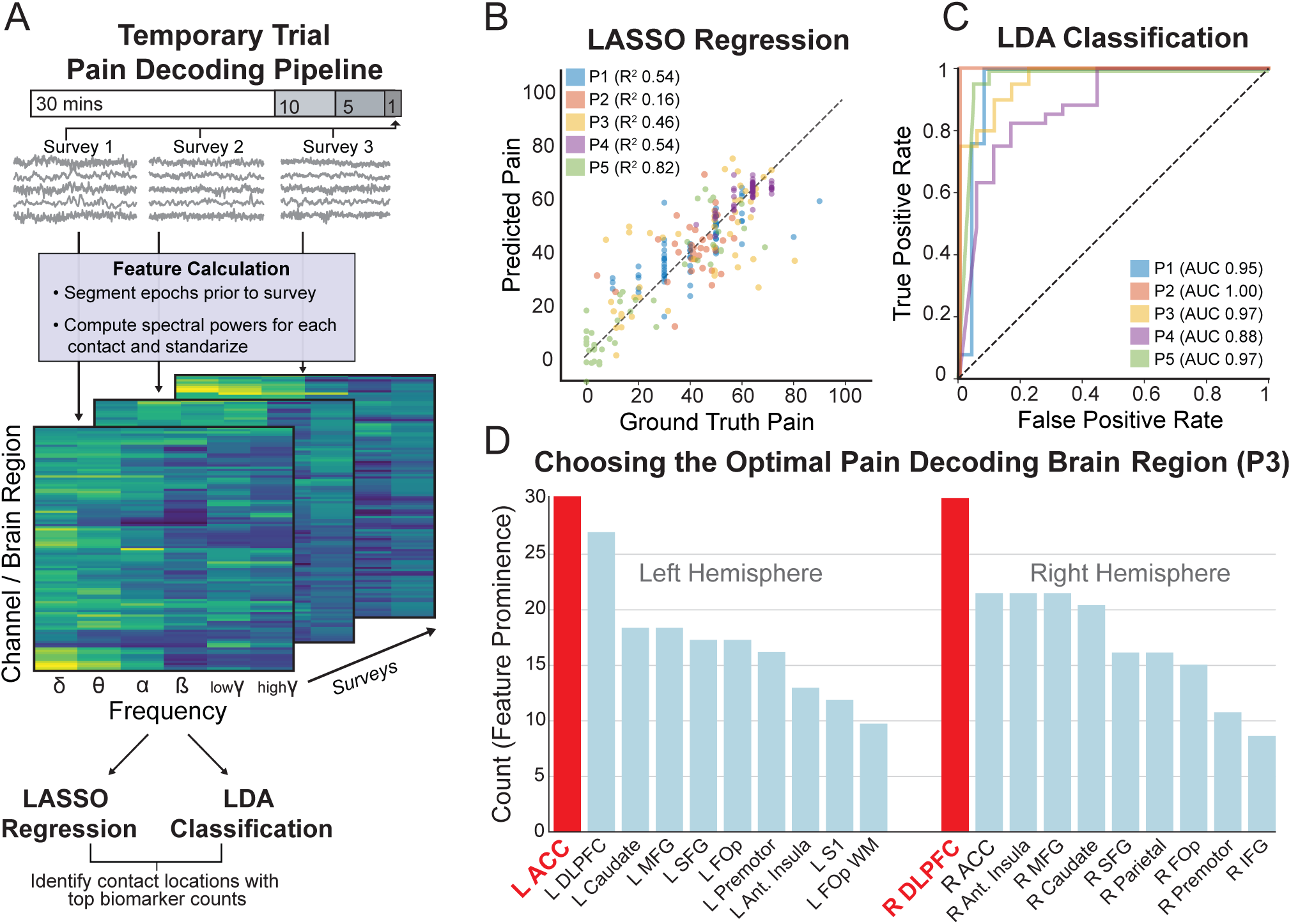
Pain decoding models identify brain regions that harbor pain biomarker signals. (A) Schematic of pain decoding pipeline applied to temporary brain stimulation trial. iEEG signals, recorded across hundreds of electrode channels across the brain, were collected from four different time epochs (1,5,10,30 mins) just prior to each pain symptom survey. iEEG signals from each epoch were used to compute spectral power within canonical frequency bands (Δ, θ, α, β, lowψ, highψ) for each electrode contact channel (colored matrix). Separate models were constructed for each time epoch using LASSO regression and LDA classification to predict 4 different pain metrics per participant (16 models per method, see Methods and Section S7). (B) Example regression models, for the best performing pain metric / time epoch per participant, show significant continuous pain prediction. (P1: NRS/5min, P2: VAS/30min, P3: VAS/10min, P4: NRS/10min, P5: VAS/10min). (C) Example receiver-operating-characteristic curves of LDA classification models for best performing pain metric / time epoch per participant also show excellent area under the curve (AUC) performance on discriminating high vs low pain. (P1: NRS/5min, P2: reliefVAS/10min, P3: VAS/5min, P4: NRS/1min, P5: VAS/10min) (D) Bar graph showing number of instances (Count) that individual brain regions appeared amongst the top 10 weighted features, across all significant regression and classification models in each hemisphere, for an example participant P3. The top brain region (red bars) was selected for permanent implant. Abbreviations as per text.

Model inputs consisted of spectral-power features (δ, θ, α, β, low γ, and high γ) from sEEG recordings at four-time durations preceding each survey (1,5,10 and 30 min). Feature vectors derived from each participant’s recordings were entered into a LASSO regression model to examine the continuous relationship between neural features and best performing pain metrics (Figure 3B). The coefficient of determination (R²) ranged from 0.16 (P2) to 0.82 (P5) for significant models. We also used a linear discriminant analysis (LDA) classifier to distinguish binary high-pain from low-pain states to provide direct translation to the RC+S device’s embedded LDA capability. The LDA (Figure 3C) classifier reliably identified high versus low pain states in five participants, with area-under-the-curve (AUC) values ranging from 0.88 (P4) to 1.00 (P2) for optimal personalized metrics (see Figure 3 caption). We could not perform biomarker decoding in P6, because their pain scores did not fluctuate over a clinically meaningful range.

By counting the most important features across significant within-participant models (Figure 3D, S5.1), we most frequently identified biomarkers in the ACC, DLPFC, and caudate, although the specific pattern of dominant brain regions varied among individual participants. Taken together, these findings support the feasibility of using a data-driven analysis of intracranial recordings to accurately decode both categorical and continuous pain states and to select personalized brain targets for sensing pain biomarkers.

### Long term outcomes of closed-loop vs sham DBS for chronic pain

Based on stimulation mapping and biomarker decoding results from the temporary trial, we implanted permanent DBS electrodes targeting one optimal stimulation and sensing target in each hemisphere for each participant (Table S1, Figure 4). Participants waited 3 weeks for electrode stabilization post-surgery before we initiated up to 3 daily streaming sessions with pain symptom reports (NRS, VAS, MPQ) at home daily. Initial data collection occurred without active stimulation to validate previously discovered pain biomarkers. We turned on DBS after receiving 100 pain reports and found that minutes of stimulation at personalized targets reproduced rapid pain relief at parameters similar to those used during the temporary trial (Tables S3-9). We then optimized closed-loop programs over the next 6 months, testing settings for up to 3 weeks at a time, before commencing double-blinded testing.

**FIGURE 4:**
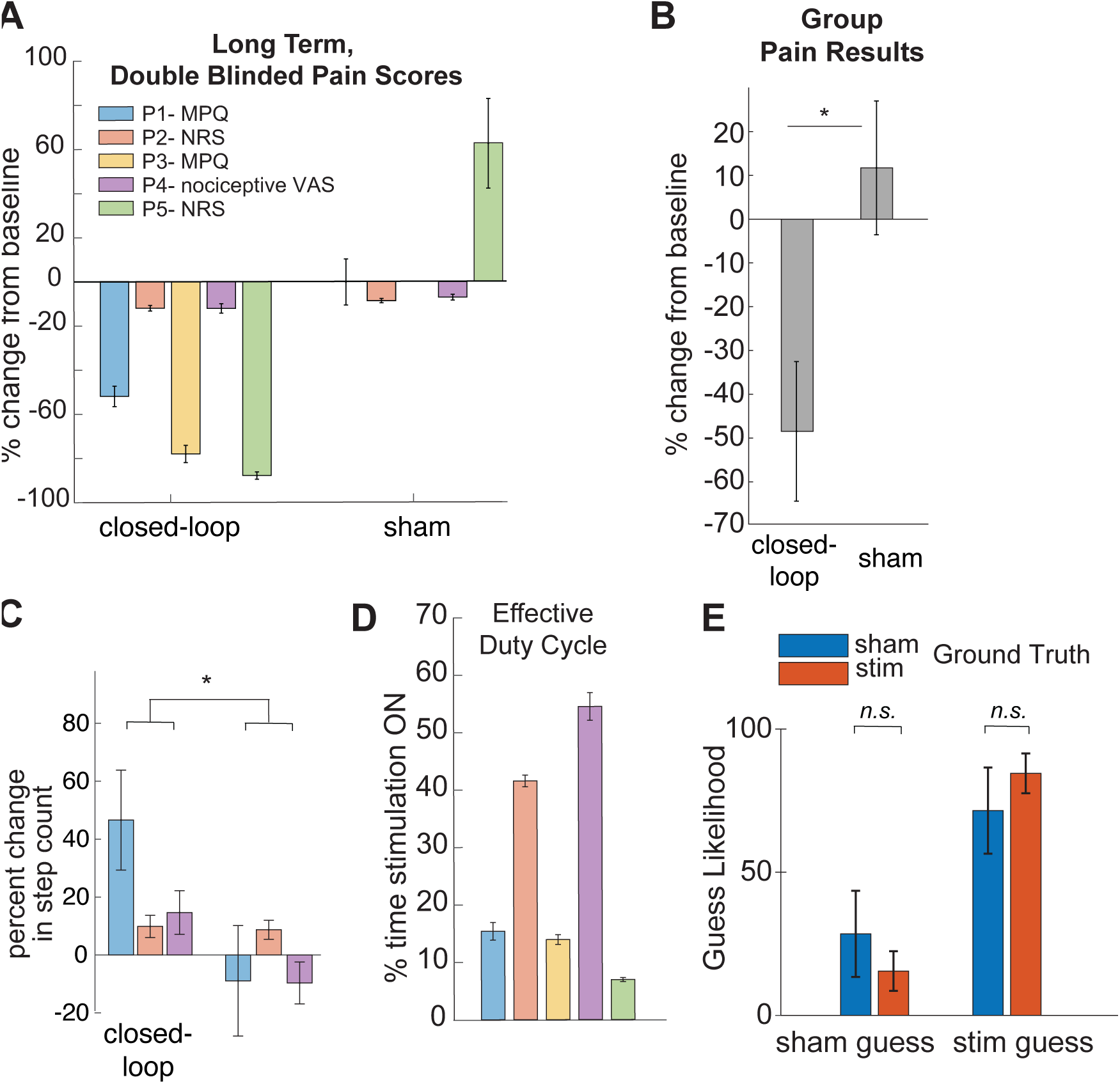
Participant and group level, long term, double-blinded outcomes demonstrate significant pain relief in response to closed-loop DBS compared to sham. (A) Participant bar plots reflect percent change in pain metrics compared to same-day, pre-stimulation baseline for closed-loop vs sham conditions. Individually representative pain metrics were chosen as per IMMPACT guidelines. (B) Group level mixed effects model showed 50% improvement of pain compared to baseline across all participants. Closed-loop DBS was significantly different from sham (see text). (C) Percent change in mean daily step count (for available data) shows a significant improvement in physical function during closed-loop DBS compared to sham in two of three participants. (D) Bar plots show average daily stimulation duty cycle during closed-loop DBS. (E) Group bar plots confirm adequate blinding, with no significant difference in stimulation guesses across groups. All error bars show standard error of the mean. *p<0.05, n.s.=not significant.

In the double-blind, crossover phase, each participant underwent randomized stimulation with either closed-loop or sham DBS (see Methods). Baseline pain scores did not differ significantly among conditions. Closed-loop DBS produced pronounced and significant reductions in chronic pain (Figure 4A, using personalized metrics) relative to baseline, whereas sham conferred minimal benefit (linear mixed effects model F_1,304_=6.43, R^2^=0.88, p=0.012). Closed-loop DBS reduced daily averaged pain intensity by approximately 50% (range 12 to 87, Figure 4B) compared to a mean pain increase of 11% with sham. A proxy for physical function, daily step count (Figure 4C), significantly improved by 18% with closed-loop DBS compared to 1% during sham in available data (t_1,203_=2.9, p=0.001). The effective duty cycle, reflected by the percentage of time DBS was on (Figure 4D), ranged from 7-55%, substantially lower compared to always-on conventional open-loop DBS. We confirmed appropriate blinding by asking participants to guess the stimulation condition just before and after each program change (Figure 4E, S7). Participants guessed stim (78±11%) more often than sham (22±11%) regardless of ground truth (*F*(1, 4) = 6.73, *p* = 0.046). However, blinding guesses did not differ during either true sham DBS (72 vs 85%, *t(* 4) = -1.01, *p* = 0.367) or true closed-loop DBS (28 vs 15%, *t(* 4) = 1.01, *p* = 0.367). Further, closed-loop DBS was associated with significantly lower symptoms of depression and pain interference, while results of global impression of change were equivocal (Figure S9). Collectively, these results support the feasibility of personalized ambulatory neuromodulation and indicate that meaningful clinical improvements in function parallel reported reductions in pain.

### Closed-Loop DBS design and optimization

Participants reported ambulatory pain metrics 2162.2 times (range 1435 - 2743) over a mean of 2.8 years (range 2.4 – 3.3y) with high correlation between simultaneously reported scores such as VAS and MPQ (Figure 5A, R=0.84, p<10^-50^). Ambulatory pain biomarkers were derived using spectral analysis of neural data (Figure 5B top) recorded during low (<30%) and high (>70%) pain metrics (Table S9). We focused on the most meaningful reported pain metric (reported by participants) for the ambulatory biomarker, which also showed the largest dynamic range within each participant. Biomarker performance was validated by assessing the frequency-dependent performance of LDA, which revealed a significant power difference at 7.5 Hz ±2.5Hz across 1050 samples in P1, for example (Figure 5B bottom and 5C, Section S7). We observed that pain biomarkers remained stable within most participants for up to 2 years, while they changed in other participants after 1 year (Section S9, Figure S7).

**FIGURE 5:**
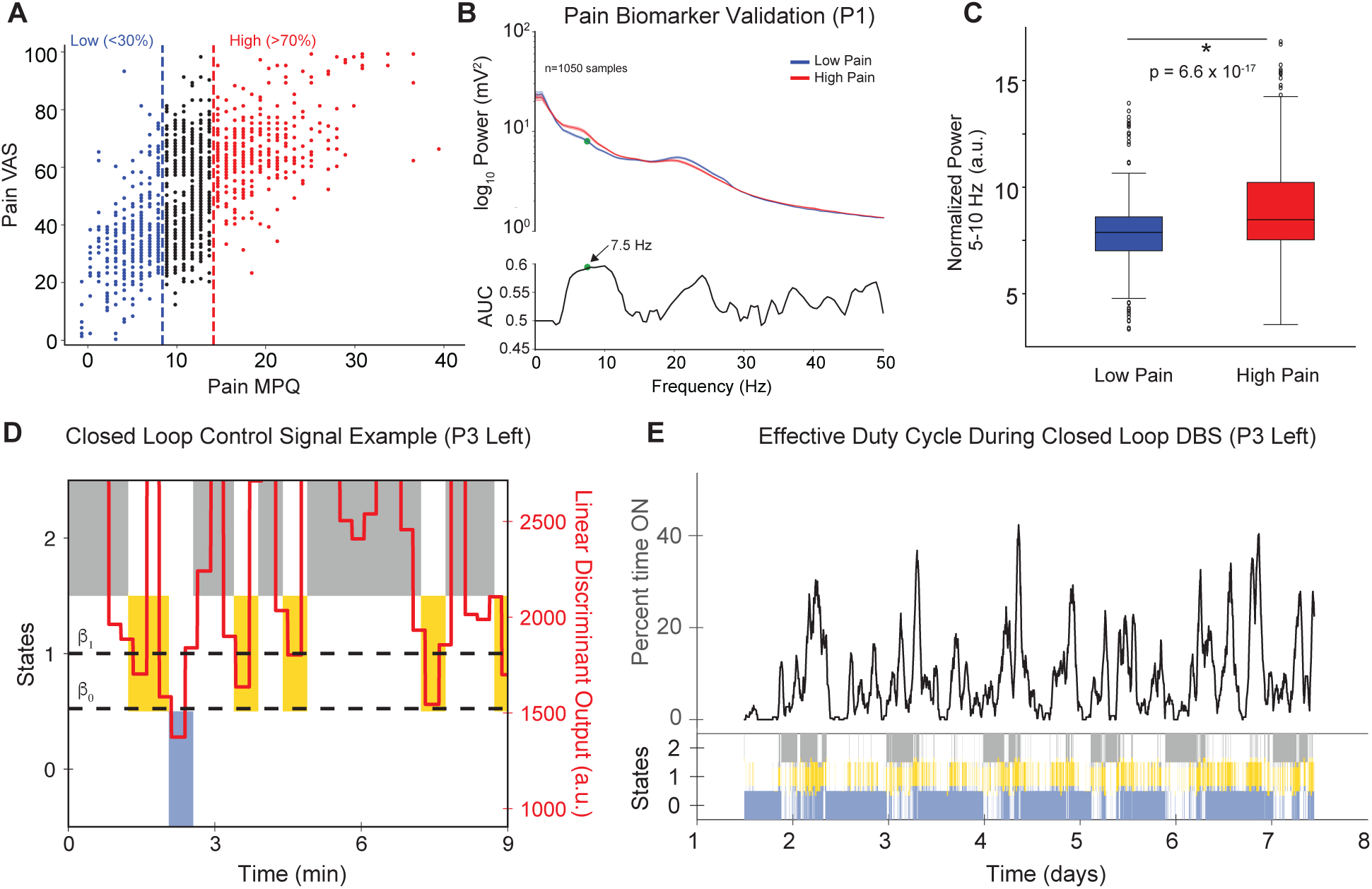
Ambulatory pain biomarker validation and closed-loop control design. (A) Pain MPQ was highly correlated with VAS, shown here for example participant P1. We dichotomized high and low pain using the upper and lower 30^th^ percentile of reports for classification to balance groups. (B) Top panel shows mean power spectra from Right ACC for all low (blue) and high pain (red) neural recordings for example participant P1. Bottom panel shows example area under the curve (AUC) performance of separate pain score classification models using all frequencies from 1-50 Hz with a 5Hz bandwidth. AUC was maximum at 7.5 Hz, indicating an optimal biomarker from 5-10 Hz for this participant. (C) Box plots show significantly higher power in 5-10Hz pain biomarker during high pain states for P1. Center line shows median, box edges show first and third quartiles, whiskers show full extent of data, and circles show outliers. (D) Nine minute example where linear discriminant output (red line, right axis) from Summit RC+S onboard classifier responded in real time to defined biomarker power inputs for P3 left hemisphere. A three state classifier, using two thresholds (β0 and β1), was used to control stimulation when LDA signal crossed thresholds for at least 30 seconds. DBS at 2 mA was delivered during state 1 only (yellow bars), while states 0 (blue) and 2 (grey) provided 0mA. (E) Example performance of closed loop controller shown in D over 7 days in same participant P3. Top panel shows percent of time that active DBS stimulation was on at home, using a 1 hour moving window. Bottom panel shows raster indicating entry events into each state (state 1 = active DBS as in D).

Panel D shows a closed-loop control signal derived from LDA distinguishing low pain states from high pain states over a nine-minute recording interval in P3 (Video S3). The LDA signal reliably transitioned between three distinct states and triggered stimulation when in state 1 (high pain state between β₀ and β₁). We measured the effective duty cycle—the proportion of time stimulation was active—in closed-loop mode (Figure 5E). The mean effective duty cycle varied but remained below 40%, demonstrating the efficiency of closed-loop DBS relative to traditional continuous stimulation. These results validate the clinical feasibility of personalized closed-loop DBS, leveraging neurophysiological biomarkers to deliver targeted modulation of pathological neural activity associated with chronic pain.

## DISCUSSION

In this study, closed-loop DBS produced clinically meaningful reductions in pain compared to sham, using rigorous blinding, with sustained efficacy after 2-3.5 years. Brain targets associated with acute pain relief during a temporary trial reproducibly provided long-term relief when permanently reimplanted. We could predict continuous chronic pain severity scores from neural power biomarkers from cortical and subcortical regions in each participant with significant accuracy. It was feasible to integrate personalized pain biomarkers into closed-loop DBS algorithms to deliver stimulation, at home, in response to ongoing predicted pain states.

Historical pain DBS cases targeting VPthal and PVG provided early enthusiasm, but many patients eventually lost analgesic effect over time due to habituation^8,27,28^. More recent efforts targeting limbic cortical regions such as ACC using continuous stimulation have similar diminishing efficacy with time^16,26^. Here we demonstrate a first attempt at implementing relatively simple closed-loop algorithms; but there is certainly room for improvement. More sophisticated feedback signals incorporating brain-wide network states or incorporating somatic, affective and cognitive pain biomarkers may provide higher efficacy. It is possible that spatiotemporal stimulation at multiple targets is required for treating multiple dimensions of chronic pain; future optimization of when and where to deliver closed-loop DBS will help to both improve efficacy and clarify mechanisms of pain processing.

Traditional pain scales, such as NRS, across patients fail to account for inter-individual variability in pain perception^29^. Consistent with IMMPACT standards for pain research^30^, we used individualized pain metrics more reflective of each patient’s personal baseline and functional goals. The apparent DBS target heterogeneity across patients can be reconciled by understanding that all stimulation targets participate in descending inhibitory pain pathways connected to the action-mode and somato-cognitive-action networks^14,31^. Effective pain relief after stimulation of ACC, medial thalamus and basal ganglia in the present study highlight likely involvement of cortico-striatal-thalamocortical loops^13^ underlying proposed pain networks. These results further support a network model of pain processing whereby modulating a single cortical, thalamic or basal ganglia node could modify ascending pain signals at the level of PVG, the rostroventral medulla or even spinal cord^14,15,32^. In participant P5, we also observed profound and durable pain relief of CPSP by targeting the ipsilateral ventral GPi, a novel DBS target, previously reported to eradicate post-herpetic neuralgia after a surgical lesion in one patient^33^. Whether detected biomarkers reflect direct modulation of these circuits or represent epiphenomena of network-level changes will require further mechanistic study. We note that most optimal biomarkers were identified at distinct brain targets than those used for stimulation, in contrast to proposed approaches for Parkinson’s Disease^34^ and major depressive disorder^35^.

It remains unclear whether biomarker-driven closed-loop DBS is necessary, as intermittent or randomly timed stimulation may be equally effective. However, our observation that effective closed-loop duty cycles less than 50% were superior to sham suggests that closed-loop DBS may prolong battery life or reduce side effects of stimulation. Future studies should test whether closed-loop control offers advantages over open-loop or simpler stimulation strategies.

The importance of stimulating gray matter (e.g., ACC, insula) versus subcortical white matter tracts remains an ongoing question in DBS^36^ and pain given widespread central sensitization and neuroplastic changes^37^. Stimulation of white matter bundles may alter network connectivity more globally^38^, whereas gray matter stimulation may modulate local synaptic activity. Parsing these effects in future studies could inform patient-specific target selection and optimize long-term outcomes.

Our study is limited by a small sample size, constraining the generalizability of these findings to other chronic pain syndromes. Although we observed initial evidence of sustained benefits up to 3.5 years, additional follow up is required to monitor for late-emerging tolerance. Future mechanistic studies are required to assess the extent to which pain biomarkers may be causal or if a combination of biomarkers can help to refine DBS control signals. The incorporation of personalized fiber tract imaging and functional MRI may help to guide personalization and further elucidate the mechanistic underpinnings of DBS-induced analgesia.

## METHODS

### STUDY OVERVIEW

The UCSF Pain Relief with Individualized brain Stimulation (PRISM) study is a single institution clinical feasibility trial using closed-loop DBS to treat refractory chronic neuropathic pain. Participants with chronic pain underwent two brain implant surgeries: first, an acute stimulation-response trial using temporary, inpatient intracranial stereoencephalography (sEEG) and electrocorticography (ECoG) electrodes to identify optimal stimulation sites and biomarkers for pain relief over 10d. Participants that obtained significant relief (Section S1-2, Table S1) were then implanted with two permanent DBS devices (Medtronic Summit RC+S) to record ambulatory neural signals and deliver on-board closed-loop stimulation in response to programmed biomarkers. This study was approved under an Investigational Device Exemption from the FDA (NCT04144972) and approved by the Committee on Human Research at the University of California, San Francisco.

### PARTICIPANTS

Six participants, aged 48-58 (3 female/3 male), with various refractory pain syndromes met inclusion criteria (Sections S1, S2) and were enrolled in the temporary brain stimulation mapping trial. A subset of five participants who obtained pain relief (Table S1) proceeded to permanent brain implant. All participants rated their baseline pain equal or greater than 6/10 on initial visit, were free of untreated psychological disorders (Table S2), and had been failed by all existing, attempted therapies. Three participants suffered from chronic post stroke pain (P1, P5, P6), one from chemotherapy induced neuropathy (P2), one with complex regional pain syndrome (P3) and another with spinal cord injury related pain (P4). Two participants (P1 and P2) experienced serious adverse events related to surgery with no serious adverse events related to stimulation (Section S1 and Figure S1).

### BRAIN IMPLANT SURGERIES

All surgeries occurred between 2019 and 2023 under general anesthesia (Section S3).

#### Temporary sEEG implant

Participants were admitted to the hospital for 10-days after surgical implant of 8 sEEG depth electrodes (PMT and ADTech) and two electrocorticography (ECoG) electrode arrays over sensory (S1) and motor (M1) cortex in each hemisphere (PMT). sEEG electrodes contained 8-14 cylindrical contacts spaced 5mm apart center-to-center terminating in the following targets: bilateral ACC, insula, subgenual cingulate, striatum, CMthal or VPThal, PVG, and globus pallidus internus (GPi) (Figure 1). Depth electrodes were stereotactically implanted using a small twist drill in the skull and secured in anchor bolts in the skull. Subdural ECoG arrays containing 8-20 contacts were inserted through a parietal burr hole to span S1 and M1 cortices. Accessible brain targets were chosen based on safety with BrainLab software. Externalized electrodes interfaced with a 256-channel amplifier connected to a system capable of electrical stimulation and recording. Testing began on postoperative day 1, and all electrodes were removed on day 10.

#### Permanent DBS implant

We used stereotactic brain surgery to implant four, 4-contact Medtronic 3387 cylindrical electrodes into two brain regions in each hemisphere: one electrode targeted the best stimulation site and the other targeted the optimal recording site. Electrodes were connected to a Medtronic Summit RC+S system, implanted in the infraclavicular location. Participants were followed weekly for up to 3.5 years.

#### Lead localization

Anatomical location of each contact was confirmed by colocalizing a postoperative CT scan with preoperative T1 MRI. We used automated parcellation on the T1 MRI image and manually verified brain regions across 13 different brain atlases in MNI ICBM template space (Section S4, Figures S2-4).

### PAIN SYMPTOM REPORTING

We collected ecological momentary assessments of pain symptoms multiple times daily using surveys on a tablet computer with REDcap v10.6.19. Surveys included pain intensity numerical rating score (NRS) and visual analog score (VAS), pain unpleasantness VAS (unpVAS), short-form McGill Pain Questionnaire (MPQ) and pain relief VAS (reliefVAS, only collected after stimulation). Pain body diagrams were completed multiple times daily using an iPad and used to qualitatively assess results of stimulation as previously described.^23^

### STIMULATION AND RECORDING SETTINGS

To determine optimal stimulation targets and parameters for each participant, we tested stimulation parameters used in prior DBS pain studies (10, 50 or 100Hz; 100-300 μs; 0-4 mA, biphasic square wave pulses)^24^. We performed exploratory iEEG mapping by delivering continuous bipolar stimulation to contact pairs in 14 predetermined targets including bilateral ACC, SGC, insula, M1 and S1, VS, thalamus, PVG and incidentally sampled regions along the path of electrodes (i.e., caudate, vmPFC) (Figure 1, Section S5, Tables S3-8). After safety testing at each site for 3s, regions of interest were stimulated in successive blocks for up to 300 seconds in pseudorandomized order with sham stimulation (0 mA). Pain reports were collected within 60s of stimulation termination. After each stimulation block, we chose up to 8 candidate regions to advance for testing in the following block based on pain surveys. Later blocks included up to 20 minutes of washout duration to allow pain scores to return to baseline. Ambulatory stimulation with the Summit RC+S device used biphasic stimulation with active recharge and sham used 0mA. Because one participant declined ambulatory sham testing (P3) we could only collect sham stimulation results in 4/5 participants.

iEEG data during the temporary trial were recorded in a similar manner to that previously described for drug-resistant epilepsy (see Section S6).^25^ For ambulatory testing, participants were instructed to initiate brain data streaming sessions at home three times daily for at least five minutes. iEEG time domain signals from Summit RC+S were sampled at 250 Hz, from four bipolar channels on both devices. Eight power domain signals were sampled at 2Hz with defined frequency ranges specific to each participant’s pain biomarkers. All signals were recorded via Bluetooth on a tablet computer and streamed to a cloud platform for offline analysis using Python v3.0.

### PAIN PREDICTION MODELING

To determine the optimal brain signals and timescales for decoding pain from the inpatient stage, we compared multiple models using four time durations of neural activity (1,5,10 or 30 mins) just preceding each pain survey across four pain metrics (VAS, NRS, unpVAS, reliefVAS, Figure 3). Model inputs used iEEG spectral power, computed with the Fast Fourier Transform, averaged within canonical frequency bands (delta, theta, alpha, beta, low-gamma, and high-gamma) in regions of interest. Epochs with power values above 3 standard deviations were excluded. Models were built using two methods: 1) LASSO regression, and 2) Linear Discriminant (LDA) classification of low (lowest 30%) vs high-pain (highest 30%) surveys with leave-one-out cross-validation, optimizing for least squares. We performed permutation testing to test model significance, by shuffling pain labels 1,000 times. Models with R^2^ and AUCs in the top 95th percentile were deemed significant. To identify brain regions that harbor the most important pain biomarkers, we counted the number of times that individual contacts appeared among the top 10 magnitude coefficients for each significant model. We then summed counts within anatomical regions and selected one contact in each hemisphere that both contained the highest count and had adjacent contacts with similar values (Section S7, Figures S5-6).

### CLOSED-LOOP PROGRAMMING

DBS was programmed using monopolar and bipolar surveys to confirm that reimplanted targets reproduced acute pain relief after stimulation with settings from the temporary trial. Tonic stimulation was initially delivered in 50% duty cycle (2 min on/2 min off) to allow for clean recordings during stimulation free periods. Participants could adjust amplitude within safety ranges to determine levels that provided the best comfort without side effects. We systematically tested different parameters in an unblinded fashion for up to 6 months in one hemisphere at a time, to assess pain metric outcomes and inform closed-loop control algorithms.

We designed closed-loop algorithms for each participant using a single power channel input and single threshold (Section S8, Figure S8), where DBS was delivered in response to threshold crossing using the Summit RC+S device. The input feature was multiplied by either +/-1, depending on if the high-pain biomarker was positive or negatively associated with pain. We set the onset duration (time-above-threshold to initiate stimulation) and offset duration (time-below-threshold to stop stimulation) based on offline simulations using participants’ available ambulatory neural data. A second LDA classifier was set based on patient-specific analysis of sleep episodes (to deactivate DBS during sleep,). We iteratively tested hemisphere specific closed-loop programs for each participant over 3 months before assessing biomarker stability (Section S9, Figure S7) and choosing control algorithms for double-blinded testing (Section S10).

### BLINDING

#### Temporary Trial

Participants and investigators (except the stimulator operator) were blinded to stimulation settings throughout and to stimulation onset in the last inpatient 3 days. Sham-control stimulation used 0mA at 100Hz to contacts outside regions of interest, with identical indicator lights and sounds as verum stimulation.

#### Permanent Implant

Participants were blinded to stimulation condition in the final 6 months of the ambulatory phase by removing stimulation details from their interface tablets. An investigator remotely updated stimulation programs weekly on participants’ devices while the tablet screen was visually confirmed closed. Participants completed surveys to assess blinding before and after each program change.

### STATISTICS

Group wise pain scores were compared using Wilcoxon’s rank sum test without correction for multiple comparisons as pre-planned. Significance of decoding models was individually tested by generating 1000 surrogate null models obtained by randomly shuffling the training pain scores or high/low class labels. Group level outcomes were assessed using linear mixed effects, repeated measures ANOVA with one-tailed post hoc t-tests and false discovery rate correction for multiple comparisons (Sections S11-12).

PS, PAS and EC were involved in the design of the clinical study. All authors were involved in the collection, analysis, and interpretation of the data and the writing of the manuscript. Medtronic Inc. donated the Summit RC+S device hardware for the permanent implants, but did not have any role in data collection, analysis, interpretation or writing of the manuscript. All authors vouch for the accuracy, ethical conduct, and completeness of the data collection and for the fidelity of the study to the protocol. Informed consent was obtained from all participants before study enrollment. PS, PAS and EC have obtained a patent related to the use of closed-loop DBS to treat chronic pain.

## Data Availability

All data produced will be available online at the OpenScience Foundation (https://osf.io/) and Github.

